# Genomic insights into an outbreak of SHV-98, TEM-1 and DHA-1 producing *Klebsiella pneumoniae* in a paediatric cardiac unit

**DOI:** 10.1101/2025.02.14.25322074

**Authors:** Alice J. Fraser, Christopher M. Parry, Beatriz Larru, Lindsay Case, Kate Ball, Caitlin Duggan, Thomas Edwards, Eva Heinz

## Abstract

This study reports the outbreak of a carbapenem-resistant *Klebsiella pneumoniae* clonal lineage in a specialist children’s hospital in Liverpool, UK. Carbapenem resistant *K. pneumoniae* is unusual in the UK, and the identification of two cases in the same ward in December 2022 raised concerns and triggered an outbreak investigation. This led to the identification of a potential transmission hotspot in the form of shared equipment, and 11 patients that were colonized or both colonized and infected by a phenotypically similar strain were identified. Multi-locus sequence typing supported a clonal lineage, and whole-genome sequencing was performed on all isolates using short-read sequencing; three isolates were furthermore long-read sequenced to confidently resolve the plasmid structure. Comparing the whole-genome sequences confirmed a clonal lineage and strongly supports the suspected nosocomial transmission, highlighting again the risk of *K. pneumoniae* by its ability to colonise a range of surfaces. Detailed analysis of the resistance determinants indicates that the carbapenem resistance was driven by a combination of different beta-lactamases that would by themselves not convey this resistance profile, a synergistic interaction is thus likely. This highlights how highly resistant strains could be mislabelled as predicted sensitive when considering genetic determinants in isolation, which would have added further complexity to this investigation as the strains would get assessed as a mismatch to the observed resistance profile, and highlights the need to study beta-lactam resistances beyond presence or absence of specific genes but also consider co-occurrence.

## Introduction

*Klebsiella pneumoniae* is a major cause of healthcare associated infections (HCAIs) globally. Whilst it can be found in healthy individuals as part of the gut microbiome, the transition to infection can result in a spectrum of illness including urinary tract infections, pneumonia and bacteraemia (1). Critically, an increasing number of *K. pneumoniae* strains causing HCAIs are also resistant to multiple classes of antimicrobials, including the commonly prescribed β-lactams (2, 3). This growing resistance complicates patient management, leading to delays in effective treatment and often necessitating the use of last-line antimicrobials which further contributes to poor clinical outcomes (4, 5) and increased healthcare costs(6).

Despite hospitals being an essential component of a robust health system, they also serve as a conduit for the evolution and transmission of *K. pneumoniae*, particularly to susceptible patient populations with compromised immune systems and complex medical needs (7, 8). Hospital environments provide unique selection pressures, promoting the survival and proliferation of resistant strains due to the extensive use of antibiotics and the proximity of colonised or infected individuals to other vulnerable patients(9).

The expansion of specific lineages that have accumulated multiple antimicrobial resistance genes (ARGs), such as those encoding extended-spectrum β-lactamases (ESBL) from the SHV, TEM, and CTX-M families, as well as AmpC enzymes and carbapenemases, is commonly associated with hospitals and healthcare settings (10). Additionally, non-carbapenemase-producing strains may carry multiple β-lactamases whose synergistic effects can produce unusual phenotypes, such as carbapenem resistance, especially when occurring in conjunction with chromosomal mutations, such as those affecting porins (11). This resistance, in the absence of typical carbapenemase genes, may not be detected by molecular diagnostics.

The spread of these ARGs is facilitated by their frequent location on mobile genetic elements (MGEs), which enables horizontal transfer. This leads to enhanced resistance as ARGs accumulate and combine with other advantageous chromosomal mutations, promoting the rapid dissemination of multidrug resistance elements among *K. pneumoniae* (10, 12). These dynamics pose a significant challenge to both the diagnosis of infections and subsequent treatment, complicating clinical management and contributing to the ongoing threat of antimicrobial resistance.

This study presents a detailed genomic investigation into a recent outbreak of antimicrobial resistant *K. pneumoniae* in the cardiac unit of a specialist children’s hospital in Liverpool, U.K. Using whole-genome sequencing (WGS) and comprehensive bioinformatics analyses, we explain the genetic underpinnings of resistance and highlight the intra-strain evolution that occurred within outbreak strains. Our findings underscore the necessity for ongoing pathogen surveillance, including genomic investigations into outbreaks, and the need for robust infection control practices.

## Results

### Outbreak detection and clinical summary

Alder Hey Children’s Hospital in Liverpool, UK is a specialist healthcare provider for over 330,000 children and young people each year. It is a regional centre for cardiology and cardiac surgery for the northwest of England and north Wales. Patients are cared for on the cardiac unit or the critical care unit according to clinical needs. Routine screening of faecal samples or swabs for multidrug resistant organsisms (MDRO – Gram negative bacteria with resistance to extended spectrum cephalosporins, carbapenems and ciprofloxacin) are conducted on admission to the critical care unit and weekly during the in-patient stay. Blood cultures are taken as required in patients with fever or suspected sepsis.

In December 2022 two infants on the cardiac ward (Patients 1 and 9) had a blood stream infection with a carbapenem resistant strain of *Klebsiella pneumoniae*. This alerted clinicians about a potential clonal outbreak. An outbreak investigation was initiated including faecal surveillance swabs for MDRO on the infants and children on the Cardiac and Critical Care Unit and a retrospective review of faecal surveillance results over the previous year. This investigation identified that between February 2022 and December 2022, a total of 11 infants and children who were inpatients on the Cardiac and Critical Care Unit (including the two initially identified) were colonized, or both colonized and infected, with an apparently similar carbapenem resistant strain of *K. pneumoniae* (figure 1A). The isolates were designated as a carbapenem resistant *Enterobacteriales* (CRE) based on resistance to ertapenem using disk diffusion antimicrobial susceptibility testing (AST). Resistance to ceftolozane/tazobactam (CTT), ciprofloxacin, tigecycline and cefiderocol was also observed (figure 1B). An additional patient had a positive faecal screening sample (Patient 4) for a carbapenem resistant *Klebsiella pneumoniae* at the time of their hospital admission, before contact with other patients. This was designated as a community-acquired isolate.

**Figure 1.**
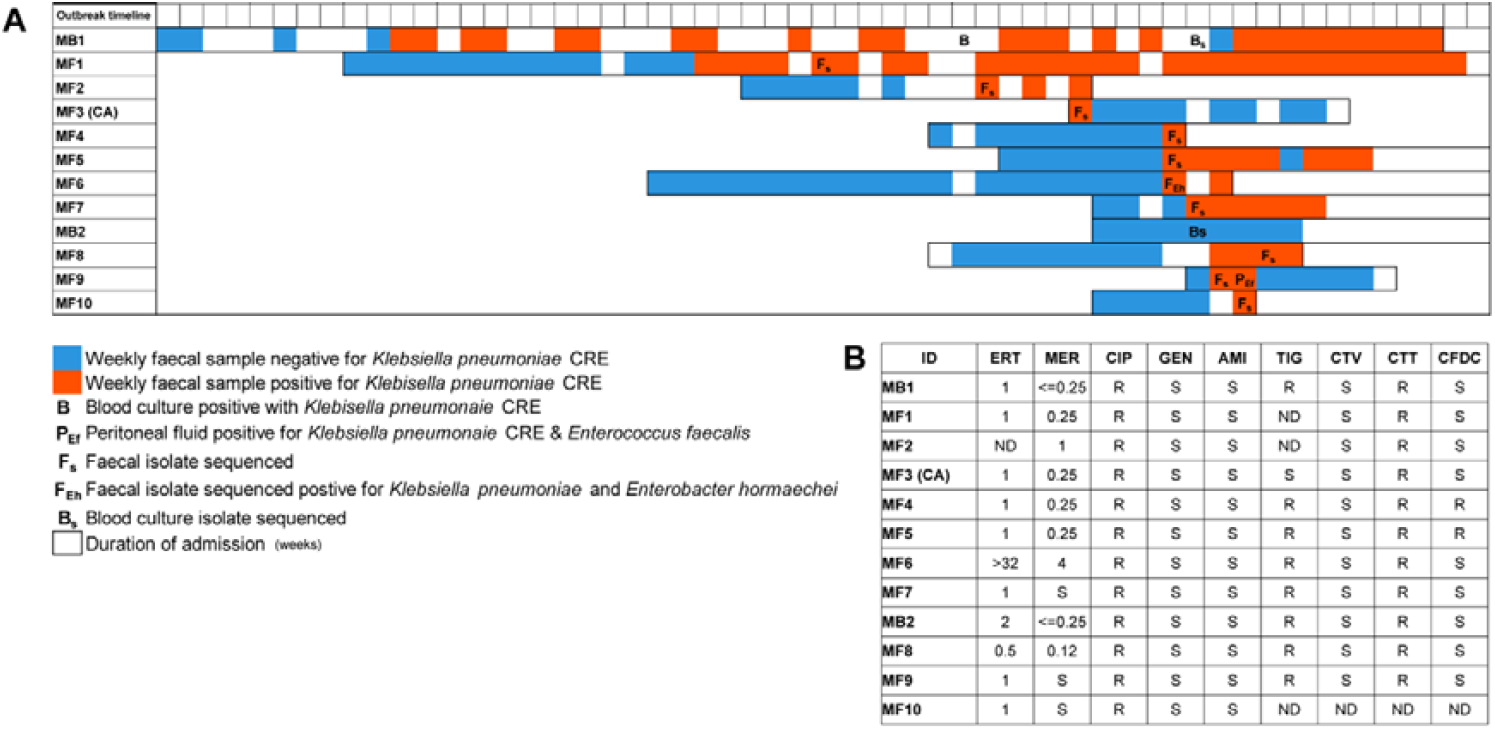
Clinical admission and sampling data for 12 patients identified as colonised and/or infected with the outbreak strain *of K. pneumoniae* **(A)**. Antimicrobial susceptibility profiles from AST performed in the hospital are also shown **(B)** (ERT: ertapenem, MER: meropenem, CIP: ciprofloxacin, GEN: gentamicin, AMI: amikacin, TIG: tigecycline, CTV: ceftazidime/avibactam, CTT: ceftolozane/tazobactam, CFDC: cefiderocol, ND: no data). The isolate from patient 7 was subsequently found to be the co-isolation of *K. pneumoniae* and *E. hormaechei*. CA indicates the community acquired isolate.

The similar antibiogram profiles of the isolates and the proximity of the patients within the same hospital ward indicated potential nosocomial transmission. This was later confirmed by multilocus sequence typing (MLST) performed by the U.K. Health Security Agency (UKHSA) at the national reference unit. Among the affected patients, nine patients were colonised with the CRE *K*.*pneumoniae* but did not exhibit signs of clinical illness, while three developed invasive infections; two of these patients (Patient 1 and 9) were bacteraemic with central line associated blood stream infections (previously mentioned), and one (Patient 11) was diagnosed with an intestinal perforation due necrotising enterocolitis leading to peritonitis with the *K*.*pneumoniae* and *Enterococcus* isolated from the peritoneal fluid. Patients with clinical illness responded to a combination of gentamicin, teicoplanin, and cefiderocol treatment (Patient 1), gentamicin, teicoplanin and ceftazidime-avibactam (Patient 9) and meropenem plus surgery for the patient with peritonitis (Patient 11).

Hospital AST using the disc diffusion method identified isolates from two patients (Patient 5 and Patient 6) as resistant to cefiderocol, according to EUCAST clinical breakpoints (zone diameter < 23 mm). However, repeat testing using the broth microdilution method subsequently determined all isolates to be clinically susceptible (MIC > 2 μg/μL) though the isolate from Patient 5 had an MIC value of 1 μg/μL which was a minimum of four times greater than the other isolates, which had MIC values or either 0.25 μg/μL or < 0.125 μg/μL.

The isolate from Patient 7 demonstrated a significantly higher MIC for ertapenem (32 μg/μL) compared to other isolates (0.5–2 μg/μL). Initial genome sequencing quality control revealed the co-isolation of *K. pneumoniae* and *Enterobacter hormachei*. Due to repeated unsuccessful attempts to culture *K. pneumoniae* in isolation, this sample was excluded from further analysis. A similar co-isolation occurred in a the peritoneal fluid sample from Patient 11 containing both *K. pneumoniae* and *Enterococcus faecalis*.

### Outbreak control measures

A number of outbreak control measures were initiated in the cardiac ward during December 2022 following recognition of the two children with bacteraemia. Screening of each patient for faecal carriage of antimicrobial resistant Gram negative bacteria at admission and then weekly during hospitalisation was routine policy on the Critical Care Unit but not the Cardiac Unit. Most children on the Cardiac Unit were already in siderooms and the ward policy mandated that those colonised with carbapenem resistant Gram negative bacteria should be isolated with contact precautions. These policies were re-inforced, with education by the Infection Control Team of staff, patients and their visitors about the detection of the resistant *K*.*pneumoniae* and how to prevent its transmission. A programme of enhanced personnel hand hygiene, personnel contact precautions, and increased environmental cleaning and decontamination was instituted. All potential medical equipment that was used between patients and that could have contributed to the cross transmission was reviewed and sent to the decontamtionation department for further cleaning. Thermometers and the scales used to weigh diapers were identified as potential contributors to the cross-transmission events as they were not always properly disinfected between patients. New thermometers and a weighing scale for each patient room were purchased. The Infection Control Team conducted daily rounds in the unit to ensure those under contact precautions were isolated and to monitor adherence to good practice among staff. Following the institution of these measures, no further patients on the ward became colonised or infected with the outbreak strain.

### Genomic characterisation of antimicrobial resistance

Initial analysis of short-read sequencing data revealed that all isolates belonged to sequence type (ST) 45 and were predicted capsule (K)-locus 62 and LPS O-antigen locus 01/02v1; all isolates were also predicted to contain an IncFIB(K) plasmid. The genomes contained several antibiotic ARGs, including *aph(3”)-lb, aph(6)-ld, bla*_DHA-1_ and the regulator *ampR*, *bla*_SHV-98_, *bla*_TEM-1_, *dfrA14, fosA, mph(A), oqxA, oqxB, qacE, qnrB4, sul1*, and *sul2*. This pattern was consistent across all isolates, except for isolate MF1-R, where *aph(3”)-lb, aph(6)-ld*, and *sul2* were not detected. Several non-synonomous chromosomal mutations were found in *acrR, ompK36* and *ompK37*, these mutations were shared between all isolates. Isolates MF2-R and MF1-R also contained the S83F mutation in GyrA. We did not identify any specific genomic determinants that explained the observed differences in resistance profiles between isolates (Figure 1B) to ertapenem, cefiderocol, and tigecycline. None of the strains were predicted to contain any genes which can induce a hypermucoid phenotype, however all isolates were positive for the yersiniabactin gene cluster, in the configuration ICEKP4/*ybt*10.

To resolve the structure and genomic location of the resistance determinants, three isolate, MB1 (from the first known colonized patient), MF3 (CA) (the community-acquired isolate), and MF9 (from the final patient admitted to the ward who later became colonized), were sequenced using both long- and short-read technologies. Assembly of isolate MB1 resulted in a circular chromosome of 5,252,750 bp, a circular IncFIB(K)/IncFII plasmid of 205,622 bp (Figure 2A), and an additional smaller plasmid of 149,366 bp classified as IncFIA(pBK30683)/IncFII(K). Additionally, two small linear contigs measuring 30,573 bp and 15,254 bp were assembled, neither of which contained any ARGs.

**Figure 2.**
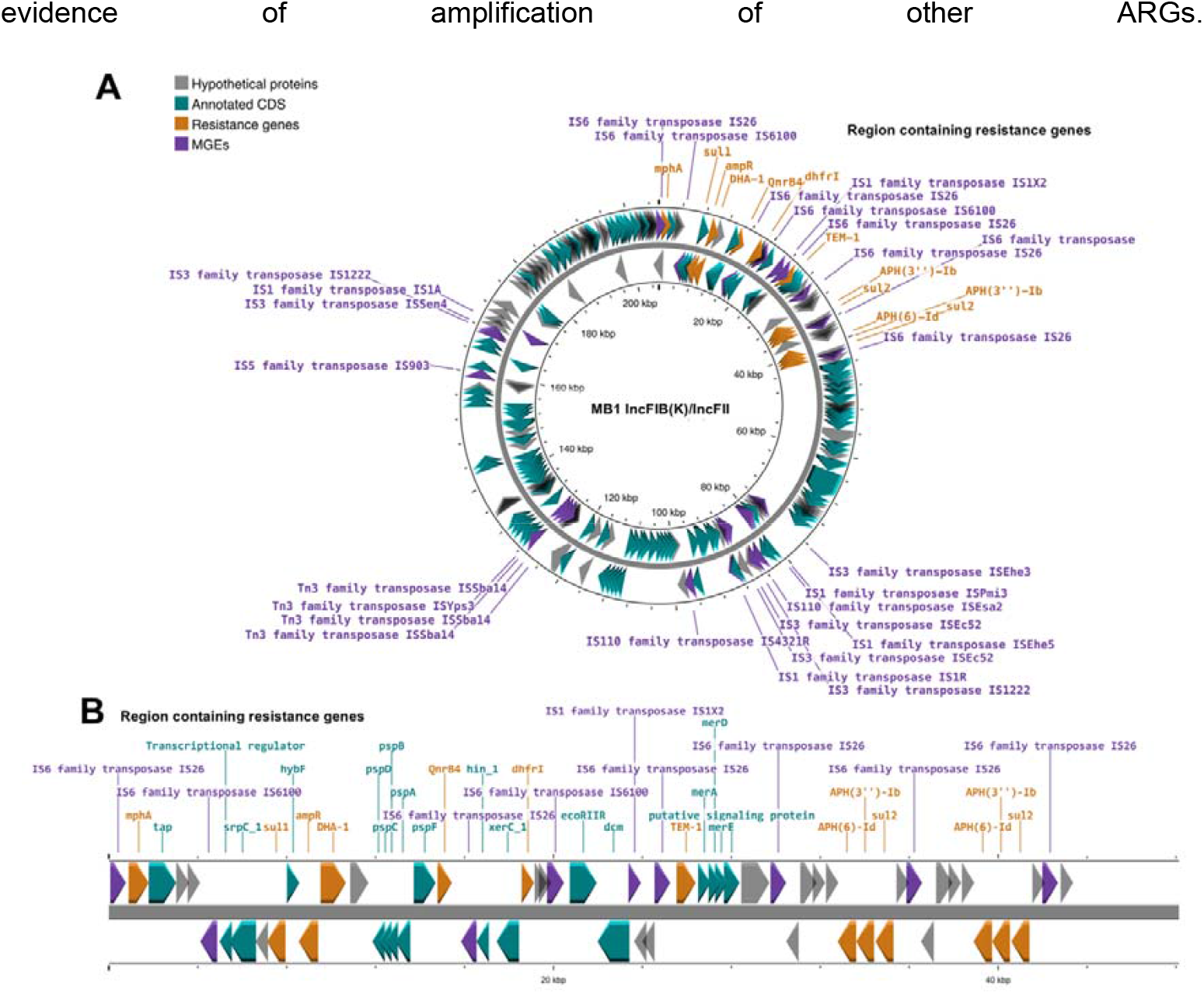
Schematic of the conjugative IncFIB(K) plasmid (A) found in all outbreak *K. pneumoniae* isolates, with genes coding for antimicro resistance-inducing proteins and mobile genetic elements (MGEs) annotated. The location of the resistance cassette containing the β-lactam genes *bla*_DHA-1_ and *bla*_TEM-1_ within the plasmid is shown along with all additional annotated coding sequences (CDS) within the resistance cass (B).

The larger IncFIB(K)/IncFII plasmid (Figure 2A) harbored multiple antibiotic resistance genes (ARGs), including the β-lactamase genes *bla*_DHA-1_ and *bla*_TEM-1_. These genes were situated within a 42,625 bp region encoding resistance genes (Figure 2B), flanked by IS*26* elements. No additional resistance genes were identified outside this region on the plasmid. However, genes linked to metal resistance and other mobile genetic elements (MGEs) were present. ARGs identified on the chromosome included *bla*_SHV-98_, *fosA, oqxA*, and *oqxB*. Apart from *aph(3’’)-Ib, aph(6)-Id*, and *sul2*, which were duplicated on the plasmid, there was no Comparisons of the genomes constructed using long-read sequencing revealed significant variations in genome size, particularly in the community-acquired isolate MF3 (CA). The total genome sizes for isolates MB1 and MF9 were 5,653,565 bp and 5,681,092 bp, respectively, while MF3 (CA) had a larger genome at 5,738,521 bp. A detailed comparison of the plasmids containing resistance genes (Figure 3) highlighted a high degree of similarity between the plasmids of MB1 and MF9, which were 205,622 bp and 205,641 bp, respectively, and both typed as IncFIB(K)/IncFII.

**Figure 3.**
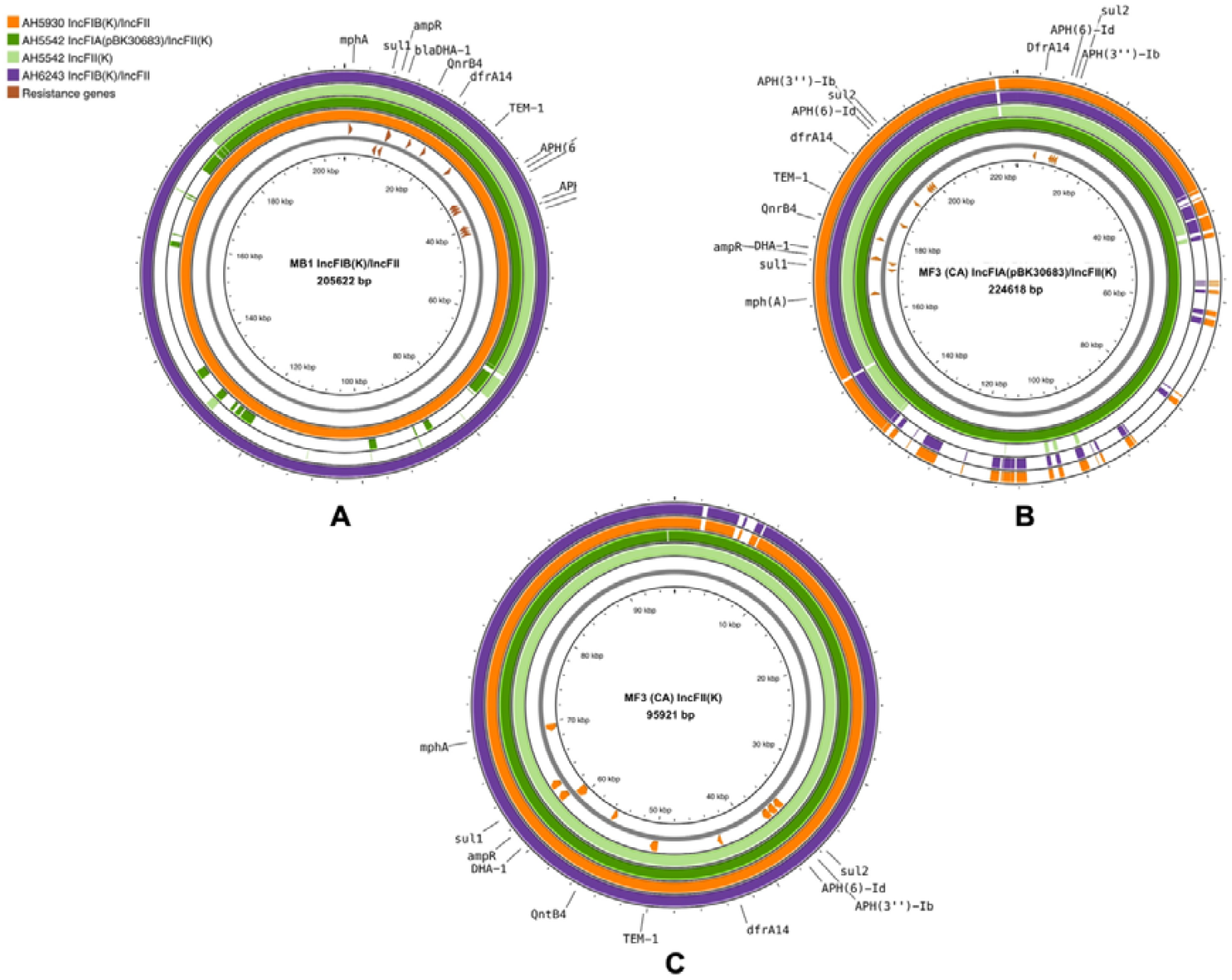
Comparative BLAST analysis of IncF plasmids from *K. pneumoniae* showing the location of ARGs. Concentric rings represent the IncFIB(K)/IncFII plasmid from isolate MB1 (from the first known colonized patient, purple), the larger IncFIA(pBK30683) plasmid from isolate MF3 (CA) (the community-acquired isolate, dark green), the smaller IncFII(K) plasmid from isolate MF3 (CA) (light green), and the IncFIB(K /IncFII plasmid from MF9 (from the final patient admitted to the ward who later became colonized, orange). **Figure A** shows all other plasmids compared to the IncFIB(K)IncFII from isolate MB1. **Figure B** shows all other plasmids compared to the IncFIA(pBK30683) plasmid from isolate MF3 (CA). **Figure C** shows all other plasmids compared to the IncFII(K) plasmid from isolate MF3 (CA). ARGs are annotated based on their position in the reference sequence, to which all other plasmid sequences were aligned.

In contrast, two plasmids harboring ARGs were identified in MF3 (CA). The larger plasmid, 224,681 bp in size, was typed as IncFIA(pBK30683)/IncFII(K), while the smaller plasmid, typed as IncFII(K), measured 95,921 bp. Both plasmids in MF3 (CA) contained all the same ARGs as those in MB1 and MF9, though the arrangement of genes varied slightly (Figures 3B and 3C). Notably, in the smaller IncFII(K) plasmid, *aph(3’’)-Ib, aph(6)-Id*, and *sul2* were not duplicated. Despite its reduced size, this smaller plasmid shared a high degree of similarity with the larger plasmid from MF3 (CA) (Figure 3C). It also resembled the plasmids from MB1 and MF9, except for a small region between 2 kb and 8 kb where sequence homology was not conserved.

The larger plasmid in MF3 (CA) contained unique regions that were absent in the plasmids from MB1 and MF9, while also lacking specific regions present in those isolates. Despite these structural differences, sequence identity was notably conserved across all plasmids in regions containing the ARGs.

To identify sub-lineages within the clonal *K. pneumoniae* outbreak collection, we employed a root-to-tip directional approach based on single nucleotide variant (SNV) distances from the ancestral node when constructing a phylogenetic tree (Figure 4) (13). Isolate MB1, obtained from the first known colonised patient formed a distinct cluster (Cluster 1) with the majority of isolates obtained from patients colonised or infected in December 2022 (MF10, MF7, MF5, MB2, MF9, and MF8), highlighting a high level of clonality and a close genomic relationship among these isolates.

**Figure 4.**
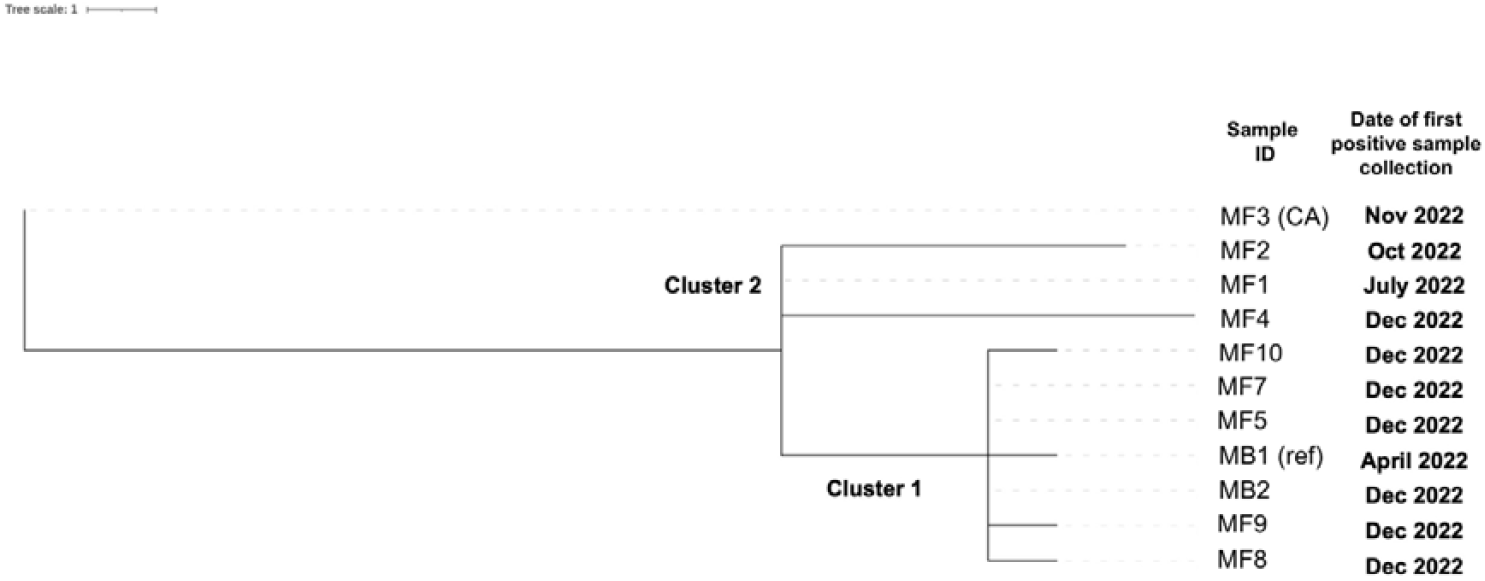
Rpincone tree constructed using a root-to-tip directional approach, according to SNV distance from the ancestral node. Isolate MB1 was used as a reference (ref). New minor lineages were defined by 5 SNVs, and new major lineages were defined by 10 SNVs, when compared to the reference. CA indicates the community acquired isolate.

Cluster 2 comprises a distinct subset of strains that are more closely related to each other than to the first collected isolate or the isolates in Cluster 1. This cluster includes isolates MF1, MF2, and MF4, which were obtained from patients colonized in July 2022, October 2022, and December 2022, respectively. The patients from whom isolates MF1 and MF2 were collected were the two earliest admissions to the ward, in March 2022 and August 2022, respectively, following the first patient (from whom isolate MB1 was obtained). Despite their temporal proximity, these isolates do not cluster with MB1.

The community-acquired isolate (MF3 CA) exhibited the greatest divergence from all other isolates, and based on our classification, it represents a separate major lineage.

## Discussion

Our investigation offers crucial genomic insights into a prolonged clonal outbreak of *K. pneumoniae* in a paediatric cardiac unit, where resistance to the carbapenem antibiotic ertapenem and the β-lactam/β-lactamase inhibitor drug ceftolozane/tazobactam was likely driven by β-lactamases SHV-98, TEM-1, and DHA-1.

Genomic analyses confirmed that all hospital-derived isolates were part of a clonal outbreak, belonged to ST45, and were of capsule type K62 and LPS O-antigen type O2a. The β-lactamase ARGs associated with ceftolozane/tazobactam and ertapenem resistance were both chromosomally-encoded (*bla*_SHV-98_) and plasmid-encoded (*bla*_DHA-1_ and *bla*_TEM-1_) and are capable of hydrolyzing β-lactam antibiotics to varying degrees. Notably, due to cumulative mutations, SHV-98 can exhibit an extended-spectrum β-lactamase (ESBL) phenotype and DHA-1, a plasmid-mediated AmpC β-lactamase, is linked to resistance against piperacillin/tazobactam (14) and carbapenems, especially when co-expressed with other β-lactamases (15) and in isolates with reduced porin expression (16), as observed in our study. The plasmid-mediated expression of *bla*_DHA-1_ is regulated by the upstream *ampR* gene (17). In our isolates, this configuration potentially leads to an expanded resistance spectrum, including resistance to ertapenem.

Despite all hospital acquired isolates being phenotypically resistant to tigecycline, we did not find any genomic determinants linked to tigecycline resistance(18). Tigecycline is typically indicated for complicated skin and soft-tissue infections, and complicated intra-abdominal infections, where other antimicrobials are not suitable(19). Although tigecycline is contraindicated in young children and infants, its off-label use has been documented in paediatric patients with infections caused by extensively drug-resistant Gram-negative bacteria (20), and it has previously been effective against ESBL-producing and AmpC-producing *Enterobacterales* (21). Understanding tigecycline resistance mechanisms in our isolates, and their interaction with β-lactam antibiotic resistance, is critical. Reporting these emerging resistance mechanisms and their association with ESBL-producing or carbapenem-resistant *Enterobacterales* is crucial for early identification and management of tigecycline resistance.

Many of the antibiotic resistance genes (ARGs) identified were plasmid-mediated and located in regions flanked by IS*26* elements, highlighting the potential for horizontal gene transfer and the further spread of resistance among *Enterobacterales*. The ARG-containing IncFIB(K)/IncFII plasmids in isolates MB1 and MF9 were highly similar. However, in the community-acquired MF3 (CA) isolate, ARGs were located on both an enlarged version of the IncFIB(K)/IncFII plasmid (typed as IncFIA(pBK30683)/IncFII(K)) and on a smaller IncFII(K) plasmid. Despite the size differences, the resistance region remained intact on both plasmids. The presence of IS*26* elements flanking the resistance region likely facilitates its mobilization and could have facilitated the transfer of the region between plasmids within the same isolate. This mechanism has been previously described for plasmid reorganization in clinical isolates with multiple ARGs (22). Furthermore, interactions between conjugative plasmids facilitated by IS elements have been identified as a general mechanism by which the transfer of ARGs is mediated (23).

Additional genomic differences were identified in the community acquired MF3 (CA) isolate when compared to the other hospital-associated isolates. Comparative analysis of core genomes revealed that the community-acquired isolate (MF3 CA) was more distantly related to the hospital-associated strains, with a 10 SNP distance to the closest hospital-acquired isolate (MF2), suggesting the *K. pneumoniae* strain is also circulating within the community.

A limitation of our study is that resistance mechanisms were inferred from genomic data with only AST used as phenotypic validation. This means we cannot definitively confirm that the observed resistance to ertapenem and ceftolozane/tazobactam was solely due to DHA-1, TEM-1, and SHV-98 production, nor can we determine if the β-lactamases acted synergistically to induce resistance. Matching unusual genotypes to phenotypic resistance is particularly important when considering the design of molecular diagnostics. While molecular CRE surveillance assays commonly screen for carbapenemase genes, our study emphasizes the importance of identifying unusual genotypes that induce carbapenem resistance, which might otherwise be missed by current diagnostics.

The outbreak’s duration and the identification of both community-acquired and healthcare-associated isolates highlight the challenges of eradicating *K. pneumoniae* strains from clinical settings. An investigation into the outbreak suggested that shared equipment may have facilitated transmission between patients and the enhancement implementation of standard infection control practices and the introduction of patient dedicated equipment subsequently halted the outbreak. Identifying phenotypic patterns, such as AST profiles, along with patient location data, is crucial for outbreak detection and management. Combining genomic investigations with phenotypic data provides a comprehensive approach to understanding the spread of resistant bacteria and mitigating further dissemination, especially in clinical settings. This approach is essential for strengthening healthcare systems and managing risks posed by multidrug-resistant organisms, particularly in vulnerable patient populations.

## Methods

### Bacterial isolates, storage and media

Clinical isolates provided on agar slants by Alder Hey Hospital, Liverpool, UK under a Material Transfer Agreement. Isolates were sub-cultured on LuriaBertani (LB) agar and then stored in glycerol broth in Microbank tubes (Pro-lab Diagnostics, U.K.) at -80 °C. For subsequent experiments, isolates were resurrected on LB agar (Sigma, U.K.) at 37 °C for 18 hours. Single colonies were then picked and used for subsequent experiments.

### Antimicrobial susceptibility testing

Initial antimicrobial susceptibility testing (AST) at Alder Hey was performed using the disk diffusion method according to EUCAST guidelines for all antibiotics except ertapenem and meropenem, where susceptibility was determined using E-tests (bioMérieux, France) according to the manufacturer’s instructions and EUCAST breakpoints. Susceptibility testing of Cefiderocol was repeated, using the broth microdilution method, according to EUCAST guidelines (24) and using cation-adjusted Mueller Hinton broth (CA-MHB) made according to Hackel *et. Al. (25)*.

### Extraction of DNA for whole genome sequencing

Single colonies were transferred to 10 ml of LB broth (Sigma, UK), and incubated overnight at 37°C, 200 RPM. Long fragments of genomic DNA, used for Oxford Nanopore Technologies (ONT) sequencing, were extracted using the Masterpure™ Complete DNA and RNA Purification Kit (Lucigen, U.K.), following the manufacturer’s instructions for the purification of DNA from cell samples. DNA used for Illumina sequencing and qPCR was extracted using the Dneasy™ Blood and Tissue Kit (Qiagen, Germany), following the protocol for Gram-negative Bacteria. The quality and size of the long-fragment DNA used for Illumina sequencing was assessed using the TapeStation (4150) system and the Genomic DNA ScreenTape Kit (Agilent, USA). All genomic DNA used for sequencing was quantified using the Qubit Fluorometer with the dsDNA BR Kit and dsDNA HS Kit (Invitrogen, USA).

### Whole genome sequencing

Short-read sequencing was performed by Microbes NG (Birmingham, UK) according to their methodology (https://microbesng.com/documents/methods/). Long-read sequencing was performed on a MinION MK1B sequencing device (ONT, U.K.). Library preparation was carried out according to the manufacturers protocol, using the ligation sequencing kit (SQK-LSK109) and Native Barcoding Expansion Kits (EXP-NBD104; all ONT). Sequencing was carried out using a FLOW-MIN106 R9.4.1 flow cell (ONT), the libraries were loaded as a pool containing a maximum of 10 sample libraries in total. Illumina and ONT reads can be found at the Sequence Read Archive bioproject ID PRJNA1211177, detailed list of accession numbers is available in Table S1.

### Assemblies

Short reads were adapter trimmed using Trimmomatic version 0.30 (26) with a sliding window quality cutoff of Q15, and assembled using Shovil (v1.1.0)(27). Basecalling and de-multiplexing of long reads was performed with Guppy (v6.4.6) (28) using the super-accurate model for basecalling. Long read sequences were then filtered using Filtlong (v0.2.1) (29) and adapters trimmed using Porechop (v0.2.4) (30). Long-read-first hybrid assemblies were produced using Flye (v2.9.3) (31), these were then visualized using Bandage (v0.8.1) (32), before being polished with long reads using Medaka (v1.8.0) (33). Assemblies then underwent polishing with short reads using Polypolish (v0.6.0) (34) and Pypolca (v0.3.0) (35). Finally, polished assemblies were quality checked, using BWA (v0.7.17) and the bwa-mem algorithm (36) and samtools (v1.9) (37) to map reads back to the consensus assembly sequence. These were then visualised using Artemis (v18.2.0) (38).

### Genome analyses

Polished assemblies were annotated using PROKKA (v.1.14.6) (37), any unannotated genes of interest were manually queried using BLAST. Sequence type, K type, O antigen group, virulence loci and chromosomal mutations were detected using Kleborate (v2.4.1) (39), with the Achtman (38) scheme for Multi-Locus Sequence Typing used. Plasmid replicons were determined using Plasmidfinder (v2.0.1) (41). Resistance genes were identified using ARIBA (v2.14.6) (40), using the SRST2 ARGANNOT data base and ABRICATE(v1.0.0) using the CARD (41) database Chromosomal mutations were identified using ResFinder (v4.1). Plasmids were visualised using Proksee (42) and compared using the inbuilt BLAST (43) tool (v1.3.1). Core genome SNPs for isolates were determined using snippy (v4.6.0) (44), using isolate MB1 as the reference. Recombinant regions were then removed from the genome alignments using Gubbins (v2.3.4) (45), polymorphic sites were removed using snp-sites (v.2.5.1) (46) and a Newick phylogenetic tree was produced using FastTree (47) and the General Time Reversible Model. The resulting tree was adjusted to SNP-length branch lengths using the python implementation of pyjar (https://github.com/simonrharris/pyjar) (48).This was then imported using Phytools (49) and further analysed using rPinecone (13); new major lineages were defined by the presence of 10 SNVs, and new minor lineages were determined by the presence of 5 SNVs. The resulting trees were visualised in iTOL(50).

## Data Availability

Illumina and ONT reads can be found at the Sequence Read Archive bioproject ID PRJNA1211177, detailed list of accession numbers is available in Table S1.

## Ethics statement

This investigation formed part of a broader study of ‘Community and nocosomial outbreaks of bacterial infections in a tertiary children’s hospital to identify different species, assess outbreak epidemiology and antimicrobial resistance’ [REC reference 19/SC/0306], South Central – Hampshire B Research Ethics Committee, Principal Investigator Professor Enitan Carrol. Details of the patient and microbiology data was extracted from the Alder Hey Hospital Meditech electronic patient record and laboratory information system by CP. It was anonymised before transfer to LSTM. Isolates were transferred to LSTM under a materials transfer agreement.

## Author contributions following CRediT taxonomy

Conceptualization – CMP, TE, EH

Data Curation – AJF, CMP, TE, EH

Formal Analysis – AJF, CD

Funding Acquisition – AJF, TE, EH

Investigation – AJF, CMP, BL, LC, KB, CD

Methodology – AJF, TE, EH

Project Administration – CMP, TE, EH

Resources – BL, LC, KB

Software – AJF, CD, EH

Supervision – CMP, TE, EH

Validation – AJF, CMP, BL, CD, TE, EH

Visualization – AJF, CMP

Writing – Original Draft Preparation – AJF, CMP, TE, EH

Writing – Review × Editing - AJF, CMP, BL, TE, EH

All authors read and approved the final manuscript version.

## Acknowledgements

We thank Professor Enitan Carrol, Alder Hey Children’s Foundation Hospital Trust and University of Liverpool for her support of this work.

